# Application of Atraumatic Care Philosophy to Children in Hospitals a Literature Review

**DOI:** 10.1101/2022.07.12.22277517

**Authors:** Reni Ilmiasih, Nurul Syafitri Ningsih

**Affiliations:** Doctoral Program, Faculty of Nursing, Airlangga University; Faculty of Nursing, Muhammadiyah Malang University

**Keywords:** atraumatic care, child-hospitalized, intervention

## Abstract

**Background:** Hospitalization for children has an impact on physical and psychological problems for children and parents. Many interventions can be done to reduce stressors for children or parents in nursing using the philosophy of atraumatic care, but there are not many articles that provide specific identification in the realm of the philosophy of atraumatic care. This study aims to determine the Evidence-Based Practice Nursing (EBPN) atraumatic care carried out by nurses in hospitals.

**Method:** This research uses the literature study method. The Literature Study stages include problem identification, searching data in 4 databases, namely Pubmed (370 Journals), Proquest (295 Journals), Clinical Key (751 Journals), and Science Direct (573 Journals) with a total of 1,989 journals after that through the screening method, assessment of study quality using the JBI Critical Appraisal Tool which resulted in the final results with 18 journals, after that through data extraction and data analysis methods.

**Results:** From 18 intervention journals to prevent physical and psychological stress, including the use of Virtual Reality (VR), and the use of buzzy and interactive play. Intervention impact separation with the presence of parents and involvement in care. Interventions related to the impact of the foreign environment are by modifying the nurse’s uniform and car orientation of the care environment. Intervention in improving treatment control with PRISM-P and Progressive Muscle Relaxation (PMR) with Guided Imagery (GI).

**Conclusion:** All interventions that are included in the 4 philosophies of atraumatic care, show the results of the effectiveness of the intervention on each principle of atraumatic care. This can be applied to patients, especially children and the elderly according to their condition.

## Background

Hospitalization for children and parents is a threatening experience (stressor), which will affect family members and family functions. These cases are often found in services for children in hospitals, which will have an impact on the psychology of children and parents(Astuti & Ilmi, 2019).

*The National Center For Health Statistics* estimates that 3-5 million children under the age of 15 are hospitalized each year. When children are hospitalized, they tend to feel abandoned by their families and feel in a foreign environment(Arifin et al., 2018). Based on a 2016 World Health Organization (WHO) survey, almost 80% of children experience hospitalization. Meanwhile, in Indonesia, based on the 2010 maternal and child health survey, it was found that of 1,425 children who experienced the impact of hospitalization and 33.2% of them experienced the impact of severe hospitalization, 41.6% the impact of moderate hospitalization, and 25.2% of the impact of mild hospitalization. (Pulungan, 2018). This shows that many children undergoing hospitalization experience moderate and severe effects of hospitalization.

Hospitalization can cause children to experience physical distress such as pain and discomfort due to injections/injections, intubation, suction, dressing changes, rectal examinations, invasive procedures, disease, immobilization, sleep disturbances, inability to drink and eat, and changes in elimination patterns(Usman, 2020). In addition to physical distress, children can also experience psychological distress such as experiencing fear, sadness, anxiety, disappointment, shame, and even anger. These circumstances can cause traumatic events in children undergoing hospitalization.

Based on research conducted byYulianawati & Mariyam (2019) on children’s reactions to hospitalization with school-age children’s anxiety levels, the results showed that there was a relationship between children’s reactions to hospitalization stressors with anxiety levels in school-age children, children’s reactions to hospitalization stressors with anxiety levels in the severe category as many as 41 people (59.4%). 36 children (52.2%), children’s reaction to the loss of control (52.2%), and 38 children’s reactions to bodily injury and pain (52.1%). Meanwhile, based on research conducted by(Gerungan, 2020)that of 30 preschool-aged children who underwent hospitalization, 16 children (53.3%) experienced moderate levels of anxiety, 11 children (36.7%) experienced mild anxiety levels, and 2 children (6.7%) experienced severe anxiety levels. and 1 child (3.3%) did not experience anxiety.

Pediatric patients urgently need access to sustainable, comprehensive, coordinated, and family-centered care(Sarjiyah et al., 2018). As child nurses or other health workers, we must have a philosophy (mindset) to be able to provide care for children by focusing on changes in the physical condition, development, and emotional needs of children. (Saputro et al., 2017).

In providing care to children, there is a philosophy of care that nurses need to pay attention to in providing nursing services, namely by focusing on the family (family-centered care) and preventing trauma (atraumatic care). (Hockenberry & Wilson, 2018). Actions are taken in dealing with children’s problems of whatever form must be based on the principle of atraumatic care or therapeutic care. Every nurse needs to understand the perspective of child nursing so that in carrying out nursing care for children always adhere to these basic principles(Wardanengsih et al., 2021).

Treatment with atraumatic care focuses on minimizing the negative impact that appears on children who are hospitalized, while family-centered care or commonly known as parental participation is a patient-centered care model and is more widely applied in the ICU, by prioritizing the participation of people. elderly in child care while in hospital(Akmalia et al., 2021). Child care requires a special method than other patients so it requires additional time in caring for children about 20%-45% of adult care time(Ministry of Health, 2020).

*Atraumatic care* is a form of therapeutic care that does not cause trauma to the child and family, whereas providing care focuses on preventing trauma and maximizing the growth and development of children in the hospital. (Kartika et al., 2021). The provision of care with atraumatic care is through actions that can reduce the physical and psychological impact, both on children and parents when they are hospitalized. Children can cry, worry, get angry, hurt and others when an event arises that can cause trauma to the child, if this is allowed it can have an impact on the child’s psychology and will hamper the child’s development. (Usman, 2020). Therefore, atraumatic care is one form of pediatric nursing in minimizing the impact of hospitalization.

Hockenberry & Wilson (2018)states that the principles that nurses must do in the philosophy of atraumatic care, include: reducing or preventing the impact of separation from the family, increasing the ability of parents to control child care, preventing or reducing injury (injury) and pain (psychological impact), modification of the physical environment. Research conducted byFeny et al. (2020)stated that the application of atraumatic care to children with hospitalization can reduce the trauma felt by children and parents. Seeing this, the nurse’s treatment of children will greatly impact the child acceptance process so it is necessary to pay attention to appropriate interventions in maximizing care for hospitalized children. If nurses can carry out their roles and functions optimally, optimal results will be obtained in meeting child care needs. The role and function of nurses who are carried out properly can minimize the negative impact of hospitalization on children.

The application of atraumatic care can minimize the psychological and physical impacts that arise from the nursing actions given, it is hoped that the child will feel safe in the hospital. Research conducted byPulungan (2018)regarding the effect of the application of atraumatic care on the anxiety response of children undergoing hospitalization concluded that the intervention group had a higher average score of anxiety before the application of atraumatic care (39.82) than the control group (37.24), while the average score of anxiety after the application of atraumatic care in the intervention group was lower (29.59) than the control group (39.71), this proves that the application of atraumatic care can reduce anxiety in children who are hospitalized. A similar thing was saidMaghfuroh (2016)in his research on atraumatic care that can reduce hospitalization anxiety in preschool children, with the result that most children (57.1%) have mild anxiety with the application of atraumatic care when hospitalized.

Nurses are expected to create nursing care with an atraumatic care approach. Atraumatic care is a form of therapeutic care provided by health workers in the child health setting, through the use of actions that can reduce physical and psychological distress experienced by children and their parents. (Maghfuroh, 2016). Actions taken dealing with children’s problems, whatever their form must be based on the principle of atraumatic care or therapeutic care because it aims as therapy for children. Atraumatic care in children can not be separated from the participation of parents. In another study conductedParulian, B., & Astarani (2018)The results obtained that hospitalization trauma prevention can be done by applying atraumatic care. The application is done to prevent injury and pain in children. Various efforts have been made by nurses to reduce physical and psychological distress in children due to the procedure.

The purpose of this research is to find out what are the implementations of Evidence-Based Practice Nursing (EBPN) atraumatic care carried out by nurses in hospitals.

## Research methods

The research design used in this study is a literature study. A literature study is comprehensive study that discusses specific related research, composed of various sources that discuss a summary of the theory and the latest findings related to the research topic. (Ridwan et al., 2021). This research is a literature study on the application of atraumatic care interventions carried out by nurses in hospitals.

Stages of the literature study as shown in Figure 1 starting from a search in four databases using keywords that are tailored to the research theme. After getting a journal that matches the keywords, the journal screening process will be continued based on the abstract, title, and full text that is adjusted to the theme of the study of literature. Furthermore, an assessment is carried out based on the feasibility of the inclusion and exclusion criteria. Stages The results of journal selection can be illustrated in the following diagram:

**Figure 1.**
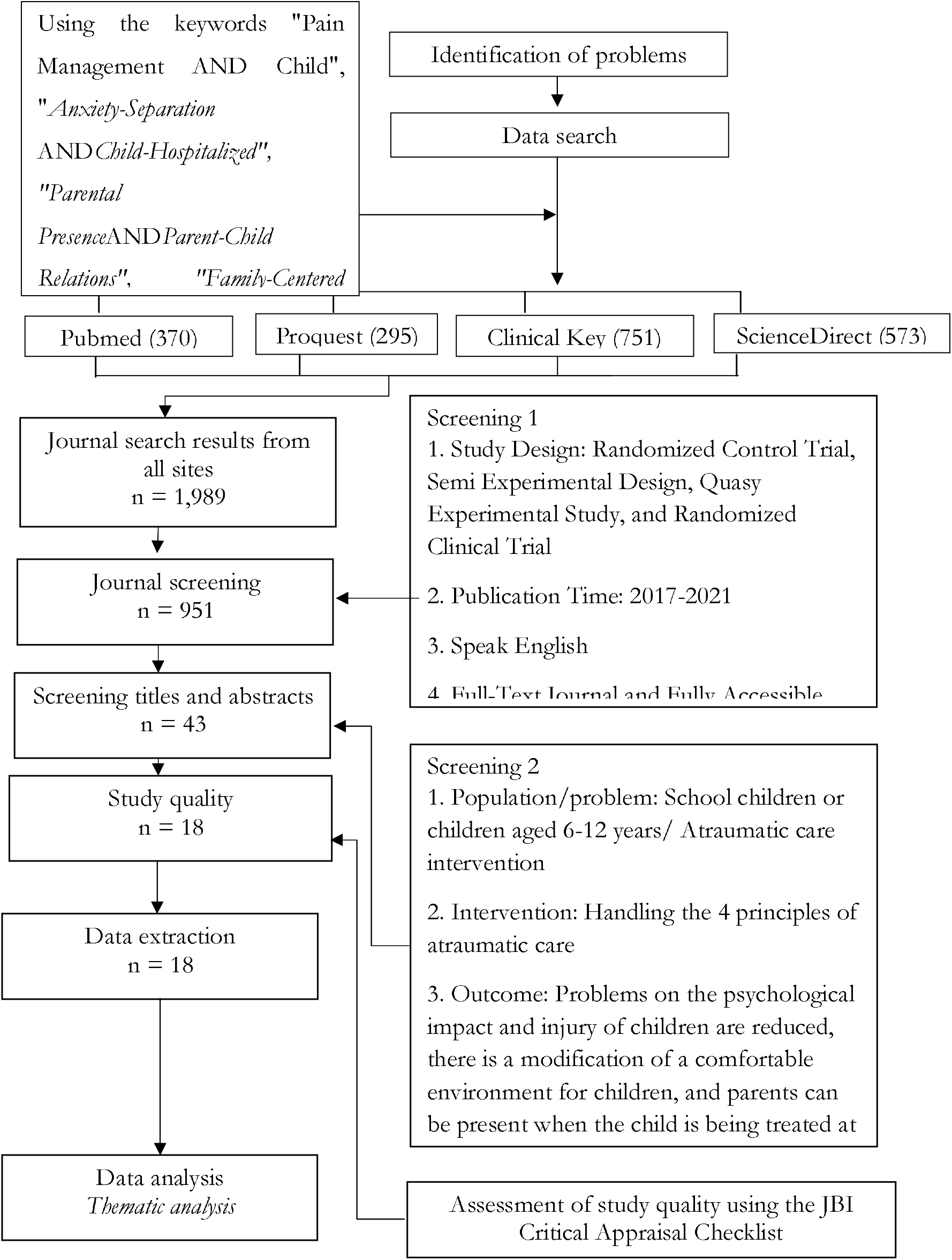
Stages of Literature Study.

### Identification of problems

Identification is the process of identifying the problem (research problem) that underlies this research to achieve the expected goals. In this literature study, researchers identified problems from various research journals as research materials. The problem raised in this study is atraumatic care intervention.

### Data search

The search for data in this literature study was carried out in several stages, including searching for articles according to the outline of the specified topic. This study uses 4 databases, namely Pubmed, EBSCO, ScienceDirect, and Biomed as sources of information. Search journals in this literature study using keywords in English, with synonym searches based on MeSH (Medical Subject Headings). The keywords in table 1 were determined using the PICO (Population/Problem, Intervention, Comparison, Outcome) method, based on the research topic “Atraumatic Care Intervention”.

**Table 1.** Keywords*Literature Review*.

|  |  |  |
| --- | --- | --- |
| ***Pain Management***<br>*Pain Management* | ***Injury***<br>*Injury* | ***Child***<br>*Child*<br>OR<br>*Child-Hospitalized* |
| ***Anxiety Management***<br>*Anxiety-Separation*<br>OR<br>*Social* | ***Anxiety***<br>*Anxiety* | ***Child***<br>*Child*<br>OR<br>*Child-Hospitalized* |
| ***Family Center Care***<br>*Clinic Outpatient* | ***Parent Role***<br>*Parent Role* | ***Parent***<br>*Parent*<br>OR<br>*Parent-Child Relations* |
| ***Parental Presence***<br>*Parental Presence* | ***Impact Separation***<br>*Impact Separation* | ***Parent***<br>*Parent*<br>OR<br>*Parent-Child Relations* |
| ***Environment Modification***<br>*Environment Modification* | ***Fear***<br>*Fear* | ***Child***<br>*Child*<br>OR<br>*Child-Hospitalized* |

### Screening

Screening is an activity to find and examine existing data and then use it as a tool to select research problems according to the research topic. In this case intervention atraumatic care, the application of intervention to the 4 principles in atraumatic care, so that the inclusion criteria are set as follows:

1. Study Design: Randomized Control Trial, Semi Experimental Design, Quasy Experimental Study, and Randomized Clinical Trial
2. Publication time 2017-2021
3. In the English language
4. Full-text journal, complete and fully accessible

Screening is carried out in two stages, namely, the first selection focuses on journal abstracts. In the second screening, the researcher specified whether the journals obtained were by the research topic.

### Study Quality

The quality of the study or the nature of eligibility in this literature study was determined by assessing the quality of the journal using the Joanna Briggs Institute (JBI) Critical Appraisal instrument. This instrument is in the form of an assessment checklist with several questions to assess the quality of each journal. This process aims to see whether there is appropriateness, alignment, and accuracy of the title, design, sample, objectives, results, and discussion. The assessment criteria are given a value of “yes”, “no”, “unclear”, or “not applicable” and each “yes” criterion is assigned 1 and the other 0. If the score is at least 50%, it is said to meet the JBI Critical Appraisal criteria, so that further review can be carried out.

### Data extraction

Furthermore, the data extraction process is carried out to find out the initial data that still meets the requirements for more in-depth analysis.

### Data analysis

After data extraction, data analysis will be carried out in the journals, which will then be processed for review in each analyzed journal, after which it will be concluded using thematic analysis which aims to obtain themes that are often discussed in research studies. Thematic Analysis is one way to review data based on identifying patterns or finding themes through data that has been collected by researchers (Heriyanto, 2018).

### Research result

#### General Characteristics

In this section, there is literature whose authenticity can be accounted for research purposes. The display of the literature results in the literature review final project contains a summary and the main results of each selected article in tabular form, then below the table section describes what is in the table in the form of meanings and trends in paragraph form. (Heriyanto, 2018).

Table 3 show that most of the research was conducted on the European continent, which was 66%. Almost half of this literature review research, 33% used the State-Trait Anxiety Scale for Children (STAIC) as the research instrument.

**Table 2.**
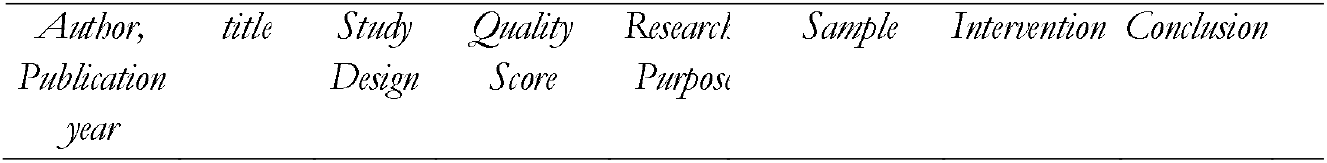
Data Extraction.

**Table 3.** General Characteristics of Study Completion

| No | Sample Characteristics | N | (%) |
| --- | --- | --- | --- |
| 1. | **Demographic Data** |  |  |
|  | Asian continent | 2 | 11% |
|  | Continent of Europe | 12 | 66% |
|  | North America | 3 | 16% |
| 2 | **Research Instruments** |  |  |
|  | Questionnaire | 2 | 11% |
|  | Yale Preoperative Anxiety Scale (mYPAS) | 2 | 11% |
|  | Pain Catastrophizing Scale for Children (PCS-C). | 1 | 0.05% |
|  | State-Trait Anxiety Scale for Children (STAIC) | 6 | 0.33% |
|  | Wong-Baker Faces Pain (WBFP) | 3 | 0.16% |
|  | Hamilton's Anxiety Scale (HAM-A) | 1 | 0.05% |
|  | Revised Children's Manifest Anxiety Scale (RCMAS) | 1 | 0.05% |
|  | Visual Analogue Scale (VAS) | 2 | 0.11% |
|  | Observational session and a parental interview | 2 | 0.11% |

Based on a literature review of table 4, from 18 journals, several interventions are following the 4 principles of atraumatic care, the first type of intervention is intervention in preventing physical and psychological trauma in children including the use of Virtual Reality (VR), the use of buzzy, the use of storytelling using books. illustrated, interactive and structured play therapy interventions, Video Glasses (VG) and interventions involving rearing animals, use of a kaleidoscope, use of aromatherapy with lavender essence, use of active and passive distraction, and use of cartoon-based endoscopic preparation packages. The second type of intervention is intervention in preventing the impact of parental separation on children including intervention that influences the presence and involvement of parents in caring for children while in hospital. The third type of intervention is intervention in modifying the environment including the use of pink and navy blue nurse uniforms and the use of toy cars for transportation of children from the inpatient room to the operating room and the fourth type of intervention is the intervention of parents’ ability to control care for children including PRISM-P intervention on parental resilience, Progressive Muscle Relaxation (PMR) and Guided Imagery (GI) intervention on parental anxiety.

**Table 4.**
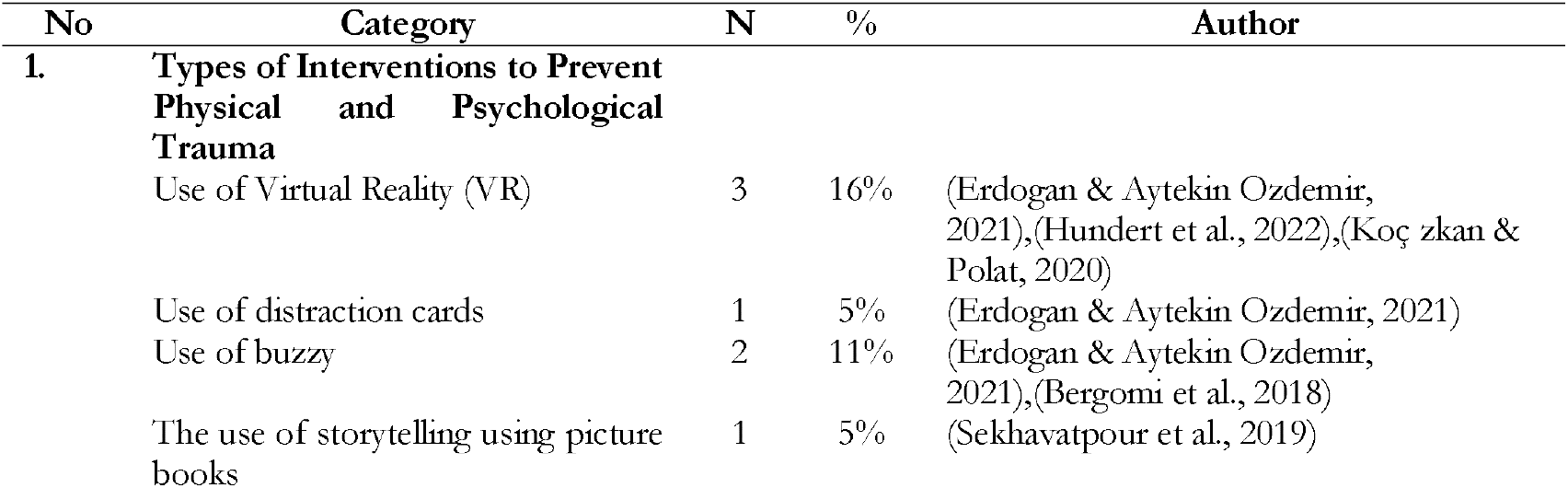
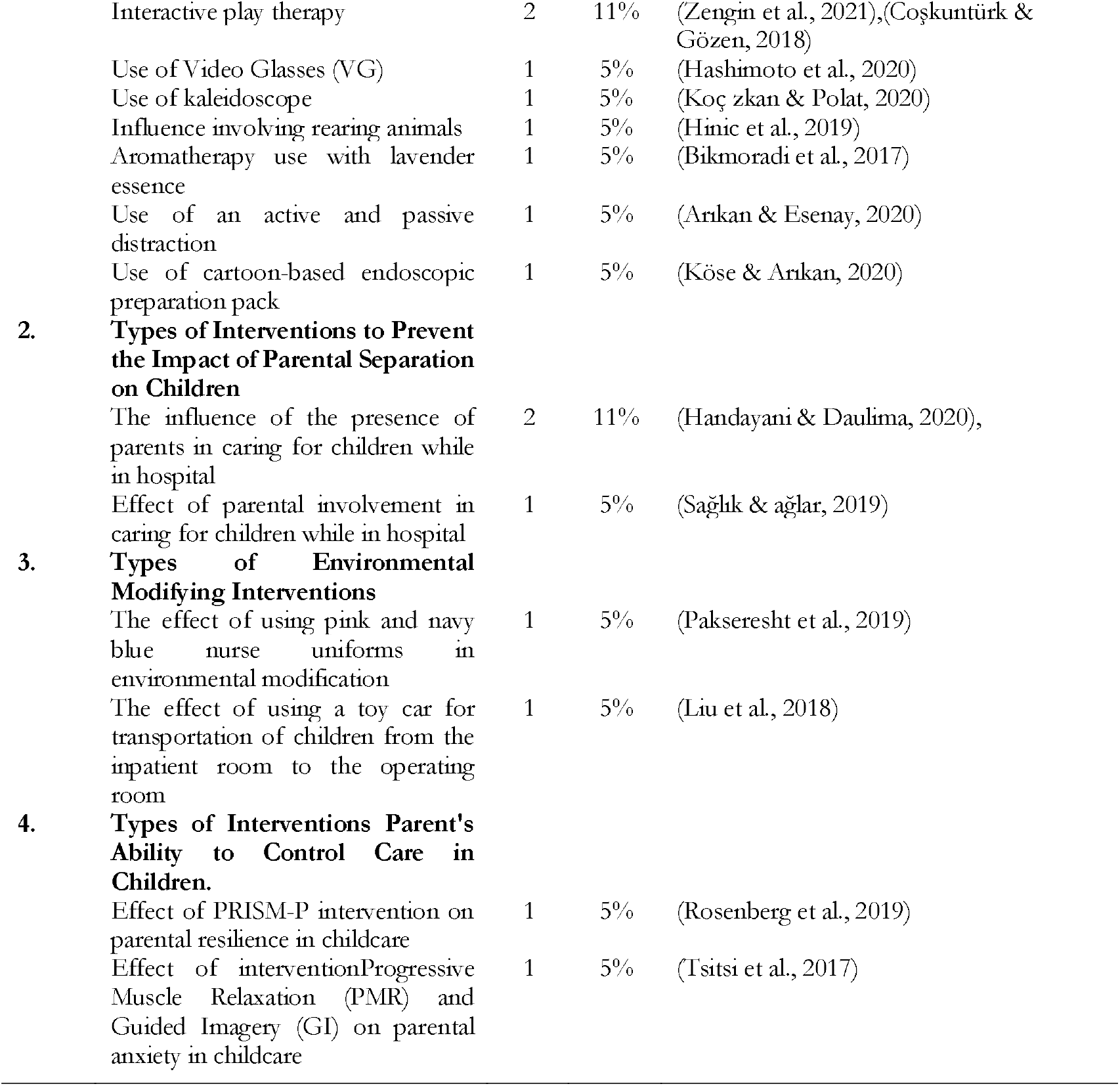
Characteristics of Interventions/Therapies on the 4 Principles of Atraumatic Care.

## Discussion

### The Effect of Types of Interventions on Preventing Physical and Psychological Trauma

Based on 18 reviewed journals, 4 types of interventions are following the 4 principles of atraumatic care for the first type of intervention, namely intervention in preventing physical and psychological trauma in children including the use of Virtual Reality (VR)has an effective effect and can be used to reduce pain and anxiety procedural venipuncture (blood sampling procedure)(Erdogan & Aytekin Ozdemir, 2021). The use of Virtual reality (VR) has been widely used in terms of reducing anxiety in children when medical procedural actions will be carried out(Koç zkan & Polat, 2020). The results in other studies also explain that the use of virtual reality (VR) is more feasible and acceptable for application in pediatric oncology for SCP needle insertion(Hundert et al., 2022). The results show that children and adolescents who use virtual reality (VR)as an intervention have more desire to continue to use virtual reality (VR)during the SCP procedure(Hundert et al., 2022).

The next intervention used of buzzy on children’s pain when taking blood samples in the group using the buzzy, the buzzy vibrator was turned on 60 seconds before the blood sampling procedure. After 60 seconds, the nurse moves the buzzy about 3 cm above the injection site and puts on a tourniquet, and performs the procedure of blood sampling. Buzzy will be active throughout the procedure. This results in the children’s overall pain increasing less in the buzzy group(Erdogan & Aytekin Ozdemir, 2021).

The next intervention is the use of storytelling using picture books on anxiety in children, The results of the study concluded that reading animated picture books were effective in reducing anxiety and behavioral disorders in children, especially after surgery. It seems that books can be a new and creative way to distract children. Telling stories before surgery can also increase the child’s familiarity with the environment, reduce the harmful effects of stress, change the child’s attitude towards the hospital environment, and surgery, and reduce the child’s fear and anxiety. (Sekhavatpour et al., 2019).

Other interventions are also carried out before surgery on children, namely using interactive and structured play therapy interventions, Video Glasses (VG), and interventions involving rearing animals in terms of reducing children’s anxiety before surgery. One study used interactive play therapy to get the results on the anxiety level of children and mothers who accompanied them before surgery, and lower levels of anxiety after surgery. It is recommended that a structured play therapy program can be used as a non-pharmacological method to reduce children’s fear of medical procedures, especially among children who have to undergo several medical procedures. (Coşkuntürk & Gözen, 2018). In-play therapyInteractive games make children role-play, where they can become doctors who will perform operations using dolls. Meanwhile, children’s structured play therapy will be tested for their ability to make something, for example, a fish figure using a ranula (cannula) without a needle injector this not only makes children’s anxiety decrease but their creativity level can also increase(Zengin et al., 2021).

On interventions of Video Glasses (VG)in the study, the results showed that there was an effective use of VG (Video Glasses) as a distraction and anxiolytic effect which was commonly used for pre-anesthesia in children. (Hashimoto et al., 2020). The findings of this study also indicated that the anxiolytic effect of the VG group was superior and resulted in lower mYPAS scores. While interventions involving pets in the study, the results of short pet therapy visits were more effective in reducing anxiety in children(Hinic et al., 2019). These results provide support for brief pet therapy with trained dogs and handlers as a means of reducing anxiety in hospitalized children. When resources to provide pet therapy visits are limited, doctors may consider prioritizing children who are most affected by anxiety(Hinic et al., 2019).

Furthermore, the use of a kaleidoscope on children’s anxiety and fear during medical procedures. A kaleidoscope is an instrument that exhibits an infinite variety of interesting geometric shapes in the shape of a flower, repeating and reflecting the image of colored glass shards on the front in a prism that reflects the inner surface. The results suggest the kaleidoscope can reduce children’s perception of pain and anxiety during venous puncture and can be used safely during painful procedures such as venous blood sampling. (Koç zkan & Polat, 2020).

Pain in children is not only when taking blood samples but one of the medical procedures, namely: intravenous catheter (injection)can make children feel pain when the needle is inserted, this makes one study using aromatherapy with lavender essence in reducing child pain when intravenous catheter(injection). The results of this study can be that aromatherapy with lavender essence helps reduce the severity of intravenous catheter insertion (injection) pain in children. Based on the results of this study as well, as well as the interest and willingness of nurses to use complementary methods, this can be applied as a feasible, safe, and inexpensive method to reduce pain severity from intravenous catheter insertion (injection). (Bikmoradi et al., 2017).

Furthermore, the intervention uses active distraction and passive distraction on the level of anxiety and fear in children during medical procedures. Active distraction referred to here is using a wooden toy that can be rotated and passive distraction using a toy bracelet to reduce pain, fear, and anxiety in children during medical procedures, namely blood sampling. Based on the research results active and passive distraction techniques are effective in reducing pain, fear, and anxiety therefore, they recommend their use during venous blood sampling. Based on the findings of this study also, playable wooden bracelets and toys are feasible distraction interventions to potentially help manage procedural pain and anxiety in children. (Arıkan & Esenay, 2020).

The last intervention that can be used to reduce children’s anxiety during endoscopic procedures is the use of a cartoon-based endoscope preparation pack. The cartoon-assisted endoscopy preparation package consists of four parts: a pillow with the character “endocan” printed on it, a cartoon, a doll, and a poster. The cast of the “endocan” character, designed by the researcher, is a 7-year-old male. The results of this study found that the cartoon-assisted endoscopic preparation package proved to be effective in reducing the fear and anxiety that may occur in children undergoing endoscopy, especially in the age group 7-12. In this case, various kinds of interventions can be done against anxiety, pain, and fear in children while in a hospital or medical action. All interventions have an effective effect on children’s anxiety, pain, and fear(Köse & Arıkan, 2020)

### The Effect of Types of Interventions in Preventing the Impact of Parental Separation on Children

Based on the 18 reviewed journals, 4 types of interventions are under the 4 principles of atraumatic care for the second type of intervention, namely intervention in preventing the impact of parental separation on children including the influence of the presence of parents in caring for children while in hospital and the influence of parental involvement in caring for children while in hospital. The intervention is one of the studies was carried out on children who were about to undergo invasive procedures such as blood collection, vascular access, intramuscular injection, and intravenous injection. Parents in the involvement group, parents were involved in invasive procedures by holding their children’s arms during blood draws, vascular access establishment, and intravenous injections, by holding their children’s feet during intramuscular injections, and by continued communication with children. them during the procedure(Sağlık & ağlar, 2019).

Parents in the parental attendance group communicated with their children during the procedure by simply being present without being involved in the procedure. The results of the study found that the pain level of children whose parents were present on the side during an invasive procedure and were involved in the procedure was lower. Given these results, we believe that parental involvement and support during invasive procedures will be effective in reducing pain levels. Meanwhile, another study explains that the presence of parents in implementing atraumatic care has a positive impact on children during hospitalization. Each hospital needs to review related policies in facilitating the presence of parents for children during hospitalization(Handayani & Daulima, 2020). This review is to support effective coping by children and meet their holistic psychological needs.

### Effect of Type of Intervention in Environmental Modification

Based on 18 reviewed journals, 4 types of interventions are under the 4 principles of atraumatic care for the third type of intervention, namely intervention in modifying the environment including the use of pink and navy blue nurse uniforms in environmental modification while reducing anxiety in children and using cars. toys for transporting children from the hospital to the operating room. Results of research use of pink and navy blue nurse uniforms found that nurses in pink uniforms produced less anxiety in children than nurses in navy blue uniforms. Providing a child-friendly environment by wearing colorful uniforms can improve the relationship between nurses and hospitalized children(Pakseresht et al., 2019).

While in research use of toy cars as a means of transporting children from the hospital to the operating room that the children who were brought to the operating room using a toy car, their anxiety was significantly reduced before pre-surgery. The ride-on toy car appears to be easy to use, highly acceptable, cost-effective, and without side effects and is a good tool for reducing preoperative anxiety in children. Both of these interventions can be used as a modification of the hospital environment for children who are being treated and undergoing medical treatment, not only making the environment safe for them but also reducing their anxiety. (Liu et al., 2018).

### The Effect of Types of Interventions on Parents’ Ability to Control Care in Children

Based on the 18 reviewed journals, 4 types of interventions are following the 4 principles of atraumatic care for the fourth type of intervention, namely the intervention of parents’ ability to control care for their children, including the PRISM-P intervention (Promoting Resilience in Stress Management Intervention for Parents)on parental resilience in child care and the effect of Progressive Muscle Relaxation (PMR) and Guided Imagery (GI) interventions on parental anxiety in child care. Children will feel safe and comfortable if their parents also feel they have the strength and are not anxious about controlling the child’s care. In this case, these two interventions refer more to the resilience and strength of parents with children who are hospitalized. The PRISM-P (Promoting Resilience in Stress Management Intervention for Parents) intervention is a short and manual intervention that targets 4 skills, namely stress management (including relaxation exercises), second goal setting (including strategies to set “SMART” (specific, measurable, actionable), realistic, and time-dependent) or (specific, measurable, actionable, realistic, and time-dependent) goals and tracking future progress, the three cognitive reframings (including skills in recognizing self-talk) and the fourth discovery benefits (including practice in identifying gratitude, meaning, and purpose despite facing difficulties) and the four benefits discoverywhichdemonstrated positive effects on parental-reported resilience and found benefit when delivered individually to parents of children with cancer(Rosenberg et al., 2019). These findings underscore an important goal in support of the PRISM-P intervention to help parents feel more resilient, which in turn can facilitate their continued ability to care for their child. While the results of the research effect of Progressive Muscle Relaxation (PMR) and Guided Imagery (GI) interventions on parental anxiety in children’s care found results of the effectiveness of the combined intervention (PMR and GI) in reducing parental anxiety. It was explained in the research that parents themselves were taught to be able to practice it themselves, considering it was easy, cheap, and did not take a long time. (Tsitsi et al., 2017). Parents play a very important role in the health outcomes of their children. So, it is very important for them to feel strong, and to support their child and all other members of the family, psychologically and physically, during this difficult time. The power of the parent itself is a confrontation with the problem.

## Closing

### Conclusion

The results of research on articles about interventions that are included in the 4 principles of atraumatic care, The first type of intervention is intervention in preventing physical and psychological trauma in children including the use of Virtual Reality (VR), the use of buzzy, the use of storytelling using picture books, interactive and structured play therapy interventions, Video Glasses (VG) and interventions involving rearing animals, use of kaleidoscope, use of aromatherapy with lavender essence, use of active and passive distraction, and use of cartoon-based endoscopic preparation packages. The second type of intervention is intervention in preventing the impact of parental separation on children including intervention that influences the presence and involvement of parents in caring for children while in hospital. The third type of intervention is intervention in modifying the environment including the use of pink and navy blue nurse uniforms and the use of toy cars for transportation of children from the inpatient room to the operating room and the fourth type of intervention is the intervention of parents’ ability to control care for children including PRISM-P intervention on parental resilience, Progressive Muscle Relaxation (PMR) and Guided Imagery (GI) intervention on parental anxiety. All interventions obtained show the results of the effectiveness of the intervention on each principle of atraumatic care. This can be applied to patients, especially children and the elderly according to their conditions.

### Suggestion

Interventions carried out for children and people are expected to be a consideration for nurses, especially those in hospital services in implementing atraumatic care interventions. From the results of this literature review, all interventions can be applied in hospitals, especially hospitals in Indonesia because they are quite safe, inexpensive, and do not take time.

## Data Availability

All data produced in the present study are available upon reasonable request to the authors

